# Serum and urine metabolomic profiles of long-term ultra-processed food intake in the longitudinal Interactive Diet and Activity Tracking in AARP (IDATA) Study

**DOI:** 10.1101/2025.02.14.25322270

**Authors:** Leila Abar, Eurídice Martínez Steele, Sang Kyu Lee, Lisa Kahle, Steven C. Moore, Eleanor Watts, Charles E. Matthews, Kirsten A. Herrick, Kevin D. Hall, Lauren E. O’Connor, Neal D. Freedman, Rashmi Sinha, Hyokyoung G. Hong, Erikka Loftfield

**Affiliations:** Division of Cancer Epidemiology and Genetics, National Cancer Institute, Rockville, Maryland, USA; Department of Nutrition, School of Public Health, University of Sao Paulo, Sao Paulo, Brazil; Division of Cancer Epidemiology and Genetics, Biostatistics Branch, NCI, Bethesda, MD, USA; Information Management Services, Inc. Beltsville, MD, USA; Division of Cancer Control and Population Sciences, National Cancer Institute, National Institutes of Health, Rockville, MD, USA; Laboratory of Biological Modeling, National Institute of Diabetes and Digestive Kidney Diseases, Bethesda, MD, USA

## Abstract

**Background:** Ultra-processed food (UPF) accounts for a majority of calories consumed in the United States, but the impact on human health remains unclear.

**Objective:** To identify poly-metabolite scores in blood and urine that are predictive of UPF intake.

**Methods:** IDATA participants (n=718) with serially collected blood and urine and up to 6 24-hour dietary recalls (ASA-24s), collected over 12-months, were selected for metabolomics analysis using ultra-high performance liquid chromatography with tandem mass spectrometry. Average daily UPF intake was estimated as percentage energy according to the Nova system. Partial Spearman correlations and Least Absolute Shrinkage and Selection Operator (LASSO) regression were used to estimate UPF-metabolite correlations and build poly-metabolite scores of UPF intake, respectively. Scores were tested post hoc in a previously conducted randomized, controlled, crossover-feeding trial of 20 domiciled participants who consumed *ad libitum* diets that were 80% and 0% energy from UPF for 2 weeks each.

**Results:** IDATA participants were 51% female, and 97% completed ≥4 ASA-24s. Mean intake was 50% energy from UPF. UPF intake was correlated with 187 (of 952) serum and 284 (of 1111) 24-hour urine metabolites (p.FDR ≤ 0.01), including lipid (n=53 serum, n=21 24-hour urine), amino acid (n=33, 59), carbohydrate (n=3, 8), xenobiotic (n=33, 69), cofactor and vitamin (n= 9, 11), peptide (n=7, 6), and nucleotide (n=6, 8) metabolites. Using LASSO regression, 28 serum and 33 24-hour urine metabolites were selected as predictors of UPF intake; biospecimen-specific scores were calculated as a linear combination of selected metabolites. Overlapping metabolites included (S)C(S)S-S-methyl cysteine sulfoxide (r_s_= –0.19, –0.23), N2-N5-diacetylornithine (r_s_= – 0.26, –0.27), pentoic acid (r_s_= –0.28, –0.31), and N6-carboxymethylysine (r_s_=0.15, 0.21). Within the cross-over feeding trial, the poly-metabolite scores discriminated, within individual, between UPF diet phases (*P* paired t-test<1×10^−5^).

**Conclusions:** Poly-metabolite scores, developed in IDATA participants with varying diets, are predictive of UPF intake and could advance epidemiological research on UPF and health.

## Introduction

The sales and consumption of ultra-processed food (UPF), defined as ready-to-eat or ready-to-heat, edible, industrially manufactured products made mostly or entirely from ingredients extracted from whole foods and often containing food substances of no or rare culinary use and/or cosmetic additives (1, 2), have increased worldwide (3). In the United States, UPF accounts for more than 50% of calories consumed by children and adults (4–6) raising concerns about the potential impact on human health. Epidemiological studies have linked UPF consumption to risk factors for chronic disease, such as weight gain and obesity (1, 7–9), as well as to risk of cardiometabolic diseases and certain types of cancer (8, 10–12).

Most epidemiological studies on UPF consumption, have used the Nova system to classify food items based on their level of processing (12). Accurate classification according to Nova necessitates detailed information on food sources, processing methods, as well as ingredients and their purposes. However, dietary assessment tools and databases capture this information to varying extents. Consequently, researchers may encounter challenges in accurately applying the Nova system, fueling concerns about exposure misclassification and comparability of study results (5, 13–16).

The integration of dietary data, measured via self-report using validated assessment tools, with biomarker measures can control confounding, minimize measurement errors, and enhance statistical power in analyses of diet and disease (17, 18). Metabolomics has been used to identify candidate biomarkers for various foods (19), beverages (20), and dietary patterns (21–23), including a dietary pattern high in UPF (5, 24, 25). Previous studies have reported on cross-sectional UPF-metabolite associations (24, 25) and one tightly controlled feeding trial (5) identified hundreds of blood and urine metabolites that differed within individual between a dietary pattern high in, compared to one void of, UPF; however, the study had a small sample size and limited menus. Currently, nutritional metabolomics research in population-based studies with longitudinal assessment of dietary intake, using validated assessment tools, and serial collection of blood and urine, for generating metabolomics data, is lacking.

The primary aims of this study were to identify serum and urine metabolites associated with average 12-month UPF intake, assessed using multiple 24-hour dietary recalls, and to develop blood and urine poly-metabolite scores predictive of UPF intake in a free-living population of participants from the IDATA study. Our secondary aim was to test whether these poly-metabolite scores could, in the context of the aforementioned, previously conducted, randomized, controlled, crossover-feeding trial, differentiate, within individual, between diets containing 80% and 0% energy from UPF.

## Methods

### The IDATA study population and design

The IDATA study was designed to evaluate the performance of web-based dietary assessment tools, including the Automated Self-Administered 24-hour Dietary Assessment Tool (ASA-24), using reference biomarkers (26). The IDATA study design and methods have been described in detail elsewhere (27). In brief, IDATA participants were recruited from a list of AARP members, aged 50-74 years, who resided in or nearby Pittsburgh, Pennsylvania, spoke English, had internet access, were not on a weight-loss diet, and were free of major medical conditions and mobility limitations. From 2012 to 2013, 1082 participants were enrolled in the IDATA study and provided biospecimens consent. The study was approved by the NCI Special Studies Institutional Review Board and is registered on ClinicalTrials.gov (Identifier: NCT03268577); all participants signed informed consent (26).

Our analytic sample included the 718 IDATA participants with dietary data, including at least one ASA-24, and available serum, 24-hour urine, and first morning void (FMV) urine, collected at two time points, 6 months apart, for metabolomics analysis (**Supplementary Figure 1**).

### Dietary data collection

Participants were divided into four groups for practical study-center related reasons and to reduce the influence of seasonal variation in diet. Each group completed the same collection activities, although the timing varied (26). Over 12 months, participants completed up to six web-based ASA-24s on a randomly assigned day approximately every other month (27, 28). Each food and beverage item reported was assigned to a unique 8-digit food code based on the foods and beverages reported in What We Eat in America (WWEIA), National Health and Nutrition Examination Survey (NHANES). The structure of the WWEIA dietary data has been described in detail elsewhere (29). Food codes were linked to the Food and Nutrient Database for Dietary Studies (FNDDS) (30), an application to convert food and beverage portions into gram amounts and to estimate nutrient values, including energy (29, 31), using standard reference codes (SR codes) from the USDA National Nutrient Database for Standard Reference.

### Nova classification for ultra-processed foods (UPF)

UPF intake was estimated according to the Nova system, which classifies foods and beverages into one of four groups based on the extent and purpose of industrial food processing. Group 1 items include unprocessed or minimally processed foods such as fresh, dried, or frozen fruits or vegetables, grains, legumes, meat, fish, and milk; group 2 items include processed culinary ingredients which are substances obtained from group 1 foods or from nature such as most plant oils, sugar, honey, and table salt; group 3 items include processed foods which are industrially manufactured foods made by adding group 2 ingredients such as salt or sugar to group 1 foods such as canned vegetables, artisanal breads, and cheese; group 4 items are UPF and include ready-to-eat foods, like store bought breads and baked goods, with ingredients not typically used in culinary preparations (2). Each ASA-24 food and beverage item was disaggregated into FNDDS food and SR codes, which were assigned to 1 of 4 Nova groups and 1 of 37 mutually exclusive food subgroups based on the WWEIA, NHANES database developed by Martínez Steele et al (29, 32). FNDDS food codes (n=44) that had not been previously assigned to a Nova group and subgroup, were reviewed and assigned a Nova classification using the “reference approach” described by Martínez Steele et al (29). Next, we calculated total energy intake per day (total calories) by summing the calories from each of the four Nova groups for each participant per recall day. We then calculated the percentage of total energy intake from UPF (i.e., Nova group 4) for each participant per recall day and averaged percentage of total energy intake from UPF across the number of ASA-24 days completed for each IDATA participant. Average, 12-month percentage of total energy intake from UPF is subsequently referred to as UPF intake.

### Biological sample collection

Participants visited the study center at months 1, 6, and 12 for assessment of anthropometry measures. During two of the center visits, either months 1 and 6 or months 6 and 12, participants had their blood drawn. Blood samples were kept at –28°C and centrifuged within two hours for 15 minutes at 3500 rpm prior to being stored at –70°C. Participants were asked to complete 24-hour urine collections at home. Urine collection days occurred approximately 7 to 10 days after a study center visit. Participants were asked to collect 100 mL of first morning void (FMV), after which they began collecting urine for the next 24-hours, including the first void of the next morning. The 24-hour and FMV urine collections were delivered to the study center by courier. The urine was weighed, aliquoted, and stored at –70°C until being sent to the biorepository where it was also stored at –70°C.

### Metabolomics analysis

Serum and urine metabolomics analyses were conducted by Metabolon Inc. using ultra-high-performance liquid chromatography (UPLC) with tandem mass spectrometry (MS/MS) to measure a broad range of metabolites including endogenously derived amino acids, carbohydrates, lipids, cofactors and vitamins, intermediates of energy metabolism as well as xenobiotics derived from exogenous sources such as food or drugs. In brief, serum and urine were analyzed in separate runs. For urine, osmolality measurements were acquired from 20 ul of each sample using a Fiske 210 Osmometer. Next, 50 ul of each urine sample was diluted (1:1) with 50 ul of ultrapure dH20 for a total volume of 100 ul. The 100 ul urine samples and all serum samples were then prepared using the automated MicroLab STAR system (Hamilton Company). Recovery standards were added, and the protein fraction was extracted with methanol, followed by vigorous shaking and centrifugation. Sample extracts were dried and reconstituted using recovery solvents containing fixed concentrations of standards. These extracts were analyzed using reverse phase/ultra-HPLC-MS/MS in positive ion mode electrospray ionization and negative ion mode electrospray ionization. Raw data were extracted, peak-identified, and processed by Metabolon Inc. using proprietary software and a biochemical reference library of >4500 known metabolites based on authentic standards. The analytical methodology used to support Metabolon’s global metabolomics platform have been described in detail elsewhere (33).

### Quality Control (QC)

Blinded, replicate, QC serum and urine samples were distributed evenly across batches at a rate of 5%. For serum, coefficients of variation (CVs) were calculated for 1464 measured metabolites using 72 replicate serum samples from 2 different pooled QC samples (n=36 QC1; n=36 QC2). The median CV for pooled QC1 and QC2 were 15.9% (IQR 8.8-30.6%), 16.2% (IQR 8.8-29.8%), respectively. CVs for 1573 urine metabolites were calculated using 144 replicate urine samples from 2 different pooled QC samples (n=72 QC1; n=72 QC2). The median CV for pooled QC1and QC2 were 18.3% (IQR 12.3-33.5%) and 18.5% (IQR 13.1-32.0%), respectively. Additionally, we included 72 duplicate samples from IDATA participants for each sample type to estimate a technical intra-class correlation coefficient (ICC) as well as a six-month, temporal ICC for each metabolite (34). Median technical ICCs were 0.89, 0.91, and 0.93 for serum, 24-hour urine, and FMV urine, respectively. Six-month, temporal ICCs were slightly lower and were more variable across the metabolites; median six-month ICCs were 0.77, 0.64, and 0.50 for serum, 24-hour urine, and FMV urine, respectively. For a given specimen type, the analysis was restricted to metabolites with <90% missingness and <30% CV.

### Statistical analysis

We used histograms to visualize the distribution of UPF consumption by sex and BMI status and tested for differences using the Kolmogorov-Smirnov test. We compared the median dietary intake of macronutrients and micronutrients, based on ASA-24s, and median concentrations of dietary reference biomarkers according to quintiles of the dietary share of UPF using Kruskal-Wallis rank sum test.

For metabolomics analysis, metabolite values below the limit of detection were assigned the minimum observed value for each metabolite and normalized by run day. To account for within-person variability in our analysis, mean metabolite levels per individual, from samples collected 6 months apart, were calculated for a given metabolite and sample type and were then natural log-transformed. We conducted a partial Spearman’s rank correlation test to identify serum, 24-hour urine, and FMV urine metabolites correlated with percentage of total energy from UPF. This analysis included 952 serum metabolites and 1111 urine metabolites. Partial correlations were adjusted for age (years), sex, race (White, non-Hispanic, African American, Asian, Hispanic), BMI category (18.5 to <25, 25 to <30, 30 to <40 kg/m^2^), and smoking status (serum cotinine detected: yes, no). To account for multiple testing, we applied a 1% false discovery rate (FDR) using Benjamini-Hochberg method such that a *p*-value of <0.01 was considered statistically significant.

To investigate associations between UPF intake and metabolic pathways, we used Fisher’s method (35) to combine *p-*values from partial Spearman’s rank correlation tests across Metabolon assigned sub-pathways. This method relies on a null distribution created from pseudo replicates, which are derived from a correlation matrix of the metabolites. The null distribution allows us to determine the significance of the combined test statistics. A FDR corrected, combined *p*-value threshold of <0.01 was considered statistically significant.

To build a poly-metabolite score predictive of UPF intake in serum, 24-hour urine, or FMV urine, we used Least Absolute Shrinkage and Selection Operator (LASSO) regression, applying 10-fold cross-validation to select the optimal λ value. To ensure the robustness of our model and prevent overfitting, we divided the dataset into training (80%) and testing (20%) sets. Using the training dataset, we performed LASSO regression to select metabolites. The prediction performance of the selected metabolites was then evaluated using the testing dataset with metrics such as mean squared error (MSE) and R-squared. Furthermore, we assessed the selection consistency of the metabolites. We applied LASSO on randomly selected 80% subsets of the dataset, repeating this procedure 100 times. The frequency of selection for each metabolite was recorded across the 100 iterations.

### UPF poly-metabolite score applied to randomized, controlled, crossover-feeding trial

We applied the UPF poly-metabolite scores from the LASSO regression models generated in the IDATA study for serum, 24-hour urine, and FMV urine samples to plasma, 24-hour urine, and spot urine metabolomics data that was previously generated in a randomized, controlled, crossover-feeding trial of 20 healthy adults (5). We tested for within individual differences in the poly-metabolite score by trial phase (80% energy from UPF for 2 weeks versus 0% energy from UPF for 2 weeks) using a paired t-test. We further considered the poly-metabolite score in a subset of 4 individuals who also participated in a second randomized, crossover-feeding trial, conducted by the same study team, where the participants consumed diets with 30% of calories from UPF with a varying percentage of calories coming from carbohydrates and fats during each 2-week phase of the 4-week randomized, crossover trial (36).

## Results

### Participant characteristics

In our analytic sample, 54% of IDATA participants were between the ages of 60 and 69 years, 51% were female, and 93% self-reported their race/ethnicity as white/non-Hispanic. There was a range of BMI, with 27%, 43% and 30% of individuals classified as normal weight, overweight, and obese, respectively. Dietary assessment completion rates were high; 97% of participants completed four or more ASA-24s (**Table 1**) and 76% completed all six.

**Table 1.** Characteristics of IDATA study participants.

|  | **Overall,** | **Female,** | **Male,** |
| --- | --- | --- | --- |
| **Characteristic** | **N = 718** | **N = 365** | **N = 353** |
| **Age** |  |  |  |
| 50-59 y | 215 (30%) | 138 (38%) | 77 (22%) |
| 60-69 y | 388 (54%) | 184 (50%) | 204 (58%) |
| ≥70 | 115 (16%) | 43 (12%) | 72 (20%) |
| **Ethnicity** |  |  |  |
| White, non-Hispanic | 667 (93%) | 327 (90%) | 340 (96%) |
| African American | 47 (6%) | 35 (9%) | 12 (3%) |
| Asian | 3 (0.4%) | 3 (0.8%) | 0 (0%) |
| Hispanic | 1 (0.1%) | 0 (0%) | 1 (0.3%) |
| **BMI** |  |  |  |
| 18.5 to <25 | 189 (27%) | 126 (35%) | 63 (18%) |
| 25 to <30 | 307 (43%) | 126 (35%) | 181 (51%) |
| 30 to <40 | 216 (30%) | 112 (31%) | 104 (29%) |
| ≥40 | 6 (0.8%) | 4 (1%) | 2 (0.6%) |
| **ASA-24s** |  |  |  |
| <4 ASA-24s | 21 (3%) | 16 (4%) | 5 (1%) |
| ≥4 ASA-24s | 697 (97%) | 349 (96%) | 348 (99%) |

Using repeated ASA-24s, collected over 12 months, the average percentage of daily calories from UPF was 51% in men, 49% in women, 49% in normal weight individuals, and 51% in overweight and obese individuals (**Supplementary Figure 2**). We also found that 12-month average macronutrient and micronutrient intake varied substantially with UPF intake. Higher UPF intake was associated with lower average energy intake from protein as well as lower 24-hour urinary nitrogen. For example, the average percentage of energy from protein in the highest compared to the lowest quintile of UPF intake was 15% and 18%, respectively. Average daily fiber density was also lowest (7.9 g/1,000 kcal) among those in the highest versus the lowest (10.7 g/1,000 kcal) quintile of average UPF intake (*p*-value<0.001). Similarly, average dietary intake of several micronutrients including vitamins A, C, D and E as well as iron, zinc, potassium, magnesium, phosphorus, and calcium were lowest among those in the highest quintile of high UPF intake (all *p-*value<0.05). Additionally, 24-hour urinary potassium concentration was lowest (*p*-value <0.001) among those in the highest quintile of UPF intake. In contrast, those in the top quintile of UPF intake tended to consume a higher percentage of energy from carbohydrates, added sugars and saturated fat (all *p-*value<0.05) and have higher concentrations of 24-hour urinary sodium (*p*-value=0.09) as compared to the individuals in the bottom quintile (**Table 2)**.

**Table 2.** Indicators of dietary content of macronutrients and micronutrients according to the dietary share of ultra-processed foods in IDATA study, using ASA-24s.

| Quintiles of dietary share of ultra-processed foods (% of total energy intake) |  |  |  |  |  |  |  |
| --- | --- | --- | --- | --- | --- | --- | --- |
|  | Overall,<br>N = 718<sup>I</sup> | Q1<br>(11.7%)<br>N = 144<sup>I</sup> | Q2<br>(11.8-42.9%)<br>N = 144<sup>I</sup> | Q3<br>(43.0-50.6%)<br>N = 144<sup>I</sup> | Q4<br>(50.7-58.2%)<br>N = 143<sup>I</sup> | Q5<br>(58.3-82.0%)<br>N = 143<sup>I</sup> | *p*-value<sup>2</sup> |
| **Micronutrient Indicators (mean density)** |  |  |  |  |  |  |  |
| **Protein** | 16.6 (14.7, 18.6) | 17.7 (15.9, 20.4) | 17.5 (15.9, 18.9) | 17.2 (15.4, 19.0) | 15.9 (14.5, 17.6) | 14.6 (12.9, 16.5) | <0.001 |
| **Carbohydrates** | 46 (42, 51) | 44 (38, 49) | 45 (40, 50) | 47 (42, 51) | 47 (43, 52) | 49 (45, 54) | <0.001 |
| **Added Sugar** | 9.2 (6.5, 13.1) | 6.3 (4.4, 8.3) | 8.4 (6.0, 10.5) | 9.2 (7.3, 12.6) | 10.9 (7.8, 14.1) | 12.9 (9.7, 16.4) | <0.001 |
| **Fat** | 35.1 (31.1, 38.9) | 34.9 (30.9, 39.2) | 35.9 (31.3, 39.0) | 35.0 (31.1, 38.6) | 34.7 (30.7, 39.4) | 35.3 (31.5, 38.5) | >0.90 |
| **Saturated Fat** | 11.7 (9.80,13.53) | 11.1 (9.29, 13.18) | 11.4 (9.78,13.34) | 11.8 (9.54,13.74) | 11.9 (10.21,13.59) | 12.0 (10.64,13.9) | 0.04 |
| **Alcohol** | 1.3 (0.0, 5.3) | 3.4 (0.0, 8.0) | 2.2 (0.0, 5.8) | 0.9 (0.0, 4.2) | 0.7 (0.0, 4.4) | 0.5 (0.0, 3.2) | <0.001 |
| **Micronutrient Indicators (mean density)** |  |  |  |  |  |  |  |
| **Fiber**<br>(g/1,000 kcal) | 9.3 (7.4, 11.6) | 10.7 (8.5, 13.4) | 9.9 (8.0, 12.1) | 9.3 (7.5, 11.6) | 8.6 (7.0, 10.7) | 7.9 (6.6, 9.8) | <0.001 |
| **Sodium**<br>(g/1,000 kcal) | 1.8 (1.57, 1.97) | 1.8 (1.57, 1.97) | 1.8 (1.61, 2.03) | 1.8 (1.62, 1.98) | 1.8 (1.56, 1.95) | 1.7 (1.53, 1.90) | 0.02 |
| **Vitamin A**<br>(µg/1,000 kcal) | 366 (287, 482) | 417 (329, 543) | 361 (302, 474) | 393 (302, 494) | 387 (271, 485) | 304 (247, 379) | <0.001 |
| **Vitamin C**<br>(mg/1,000 kcal) | 44 (29, 64) | 61 (44, 82) | 48 (35, 67) | 42 (28, 63) | 39 (26, 54) | 34 (22, 49) | <0.001 |
| **Vitamin D**<br>(µg/1,000 kcal) | 2.1 (1.41, 3.04) | 2.5 (1.61, 3.86) | 2.2 (1.48, 3.33) | 2.0 (1.56, 2.95) | 2.0 (1.29, 2.92) | 1.7 (1.18, 2.44) | <0.001 |

**Quintiles of dietary share of ultra-processed foods (% of total energy intake)**
|  | **Overall,**<br>N = 718<sup>1</sup> | **Q1**<br>**(11.7%)**<br>N = 144<sup>1</sup> | **Q2**<br>**(11.8-42.9%)**<br>N = 144<sup>1</sup> | **Q3**<br>**(43.0-50.6%)**<br>N = 144<sup>1</sup> | **Q4**<br>**(50.7-58.2%)**<br>N = 143<sup>1</sup> | **Q5**<br>**(58.3-82.0%)**<br>N = 143<sup>1</sup> | **p-value**<sup>2</sup> |
| --- | --- | --- | --- | --- | --- | --- | --- |
| **Vitamin E**<br>(mg/1,000 kcal) | 3.9 (3.24, 5.06) | 4.6 (3.58, 6.35) | 4.1 (3.39, 5.31) | 4.0 (3.24, 4.74) | 3.8 (3.17, 4.62) | 3.4 (2.97, 4.28) | <0.001 |
| **Iron**<br>mg/1,000 kcal) | 7.5 (6.47, 8.89) | 7.6 (6.49, 8.70) | 7.7 (6.62, 9.11) | 7.5 (6.54, 8.51) | 7.9 (6.50, 9.53) | 7.3 (6.18, 8.59) | 0.03 |
| **Zinc**<br>(mg/1,000 kcal) | 6.0 (5.20, 7.11) | 6.2 (5.45, 7.64) | 6.4 (5.41, 7.54) | 6.2 (5.38, 7.08) | 5.9 (5.07, 7.18) | 5.4 (4.64, 6.18) | <0.001 |
| **Potassium**<br>(g/1,000 kcal) | 1.4 (1.26, 1.67) | 1.7 (1.49, 1.85) | 1.5 (1.34, 1.70) | 1.4 (1.31, 1.66) | 1.4 (1.20, 1.54) | 1.2 (1.07, 1.38) | <0.001 |
| **Phosphorus**<br>(mg/1,000 kcal) | 676 (609, 760) | 724 (653, 808) | 708 (645, 779) | 696 (640, 765) | 665 (584, 717) | 608 (555, 675) | <0.001 |
| **Magnesium**<br>(mg/1,000 kcal) | 157 (135, 184) | 180 (157, 216) | 168 (144, 197) | 159 (142, 181) | 148 (130, 171) | 132 (119, 152) | <0.001 |
| **Calcium**<br>(mg/1,000 kcal) | 457 (375, 557) | 481 (389, 582) | 466 (391, 570) | 472 (384, 545) | 445 (370, 573) | 415 (337, 497) | <0.001 |
| **Reference Biomarkers** |  |  |  |  |  |  |  |
| **Urinary sodium** | 3780 (2801,5298) | 3625(2573,5139) | 4047 (3092,5556) | 3938 (2883,5522) | 4003 (2723,5296) | 3802 (2958,5107) | 0.09 |
| **Urinary potassium** | 3186 (2382,4188) | 3384 (2720,4477) | 3584 (2685,4502) | 3335(2471,4351) | 3052 (2233,3845) | 2762 (2031, 3594) | <0.001 |
| **Urinary nitrogen** | 89 (68,112) | 90 (70,109) | 96 (73,123) | 91 (68,113) | 87 (69,109) | 78 (63,104) | 0.004 |

<sup>1</sup> Median (IQR) /<sup>2</sup> Kruskal-Wallis rank sum test; Total percentage of energy from UPF intake were averaged across the days using multiple ASA-24s.

### Metabolite correlations with UPF intake

#### Serum

After FDR correction, mean levels of 187 (150 known and 37 unknown compounds) out of 952 serum metabolites were statistically significantly correlated with UPF intake (**Supplementary Table 1**). The 150 named metabolites included lipids (*n* =53), amino acids (*n* =33), carbohydrates (*n*=3), energy metabolites (n=4), xenobiotics (*n* =33), cofactors and vitamins (n=9), peptides (n=7), nucleotides (n=6), and partially characterized molecules (n=2) (**Supplementary Table 1**). **Figure 1A** shows the correlations between the serum metabolites that were most strongly correlated (r ≥0.20 & r ≤-0.20, p.FDR ≤0.01) with UPF intake. Serum metabolites that were positively correlated with UPF intake include: three cofactors and vitamins (i.e., carotene diol 1 & 2, and beta-cryptoxanthin), five amino acids (i.e., pipecolate, (S)C(S)S-S-methyl cysteine sulfoxide, N2-N5-diacetylornithine, 2-aminobutyrate, and gentisic acid); seven xenobiotics (i.e., ergothioneine, ethyl beta-d-glucopyranoside, 4-allylphenol sulfate, methyl beta-d-glucopyranoside, (R)-2,3-dihydroxy-isovalerate, sulfate of piperine metabolite C18H21NO3 (3), and N-acetylalliin); one partially characterized molecule (i.e., pentoic acid), and two lipids (i.e., 4-deoxythreonic acid and PE(P-16:0/22:6)). Metabolites that were negatively correlated with UPF intake include: two cofactors and vitamin metabolites (i.e., gamma-CEHC and delta-CEHC); one nucleotide (i.e., N1-methylinosine); and three lipids (i.e., O-Adipoylcarnitine, 9-decenoylcarnitine, and sphingomyelin [d18:1/18:0]).

**Figure 1.**
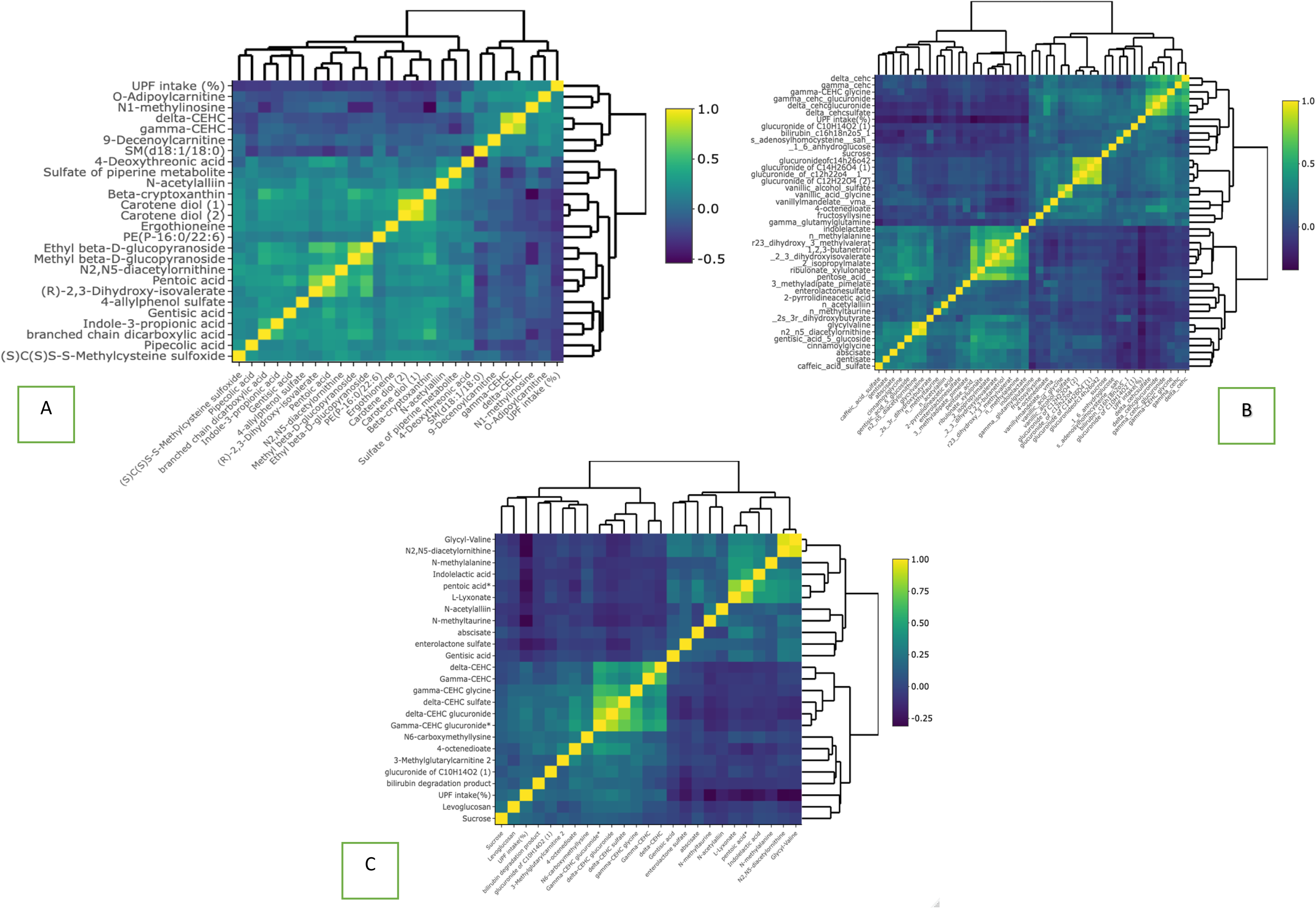
Heatmap of Spearman rank moderate significant correlation coefficients for UPF–related metabolites in serum (A), 24-h urine (B) and FMV urine. **(C)** The heatmap plot shows the correlation between UPF intake (% energy) and metabolites in 3 biospecimens. Correlations estimated using Spearman partial correlation, adjusted for age (continuous), sex (men, women), smoking (cotinine detected: yes, no), race (White, non-Hispanic, African American, Asian, Hispanic) and BMI (18.5 to <25, 25 to <30, 30 to <40 kg/m^2^). Multiple testing was corrected for using the Benjamini-Hochberg method; statistical significance was defined as a corrected *p*-value of <0.01. The dark blue color shows strong negative correlations and the light yellow color shows strong positive correlations.The dendogram uses heirarchical clustering and shows that metabolites generally cluster according the direction of the correlation UPF intake.

#### Urine

After FDR correction, mean levels of 284 (207 known and 77 unknown) and 234 (168 known and 66 unknown) out of 1111 metabolites were significantly correlated with UPF intake in 24-hour and FMV urine, respectively, with 203 metabolite correlations common to both urine types. The named metabolites in 24-hour and FMV urine include lipids (n =21 & 22, respectively; n overlapping =17), amino acids (n =59 & 47; n overlapping = 42), carbohydrates (n= 8 & 7; n overlapping = 7), energy metabolites (n=2 &1; n overlapping = 1), xenobiotics (n =69 & 51; n overlapping = 46), cofactors and vitamins (n=11 & 12; n overlapping = 11), peptides (n=6 & 5; n overlapping = 4), nucleotides (n=8 & 6; n overlapping = 4), and partially characterized molecules (n=23 & 17; n overlapping = 13) (**Supplementary Table 2-3**). **Figure 1B-C** shows the correlation between urine metabolites that were most strongly correlated (r ≥0.20 & r ≤-0.20, p.FDR ≤0.01) with UPF intake. Urine metabolites that were positively correlated with UPF intake include: one lipid (i.e., O-Adipoylcarnitine); and two cofactors and vitamin metabolites (i.e., gamma-CEHC and delta-CEHC). Urine metabolites that were negatively correlated with UPF intake include: one partially characterized molecule (i.e., pentoic acid); two amino acids (i.e., (S)C(S)S-S-methyl cysteine sulfoxide and N2-N5-diacetylornithine), and three xenobiotics (i.e., genistein sulfate, N-acetylalliin and glucuronide of piperine metabolite C17H21NO3 [3]).

#### Serum and urine

Overall, 414 metabolites were common to serum (out of 952 metabolites) and urine (out of 1111 metabolites). The UpSet plot (**Figure 2)** shows that the largest intersection in UPF intake-metabolite correlations (n=155) was for 24-hour and FMV urine. Overall, 48 UPF intake-metabolite correlations were observed in all 3 biospecimen types while 8 were common to serum and 24-hour urine, and 4 were common to serum and FMV urine. Notably, 127, 73, and 27 UPF-intake-metabolite correlations were unique to serum, 24-hour urine, and FMV urine, respectively.

**Figure 2.**
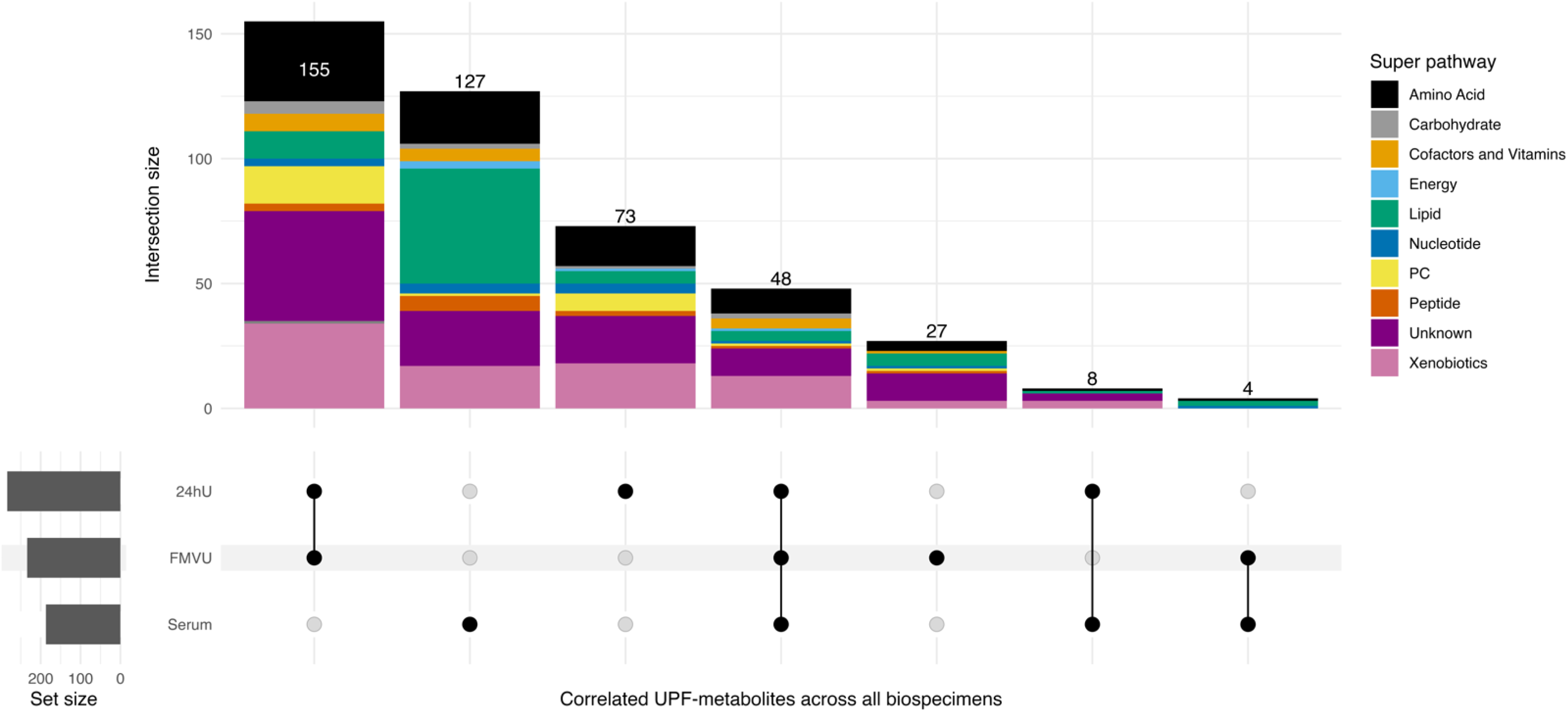
Instersection of metabolites correlated with UPF intake and their related pathways across in serum and urine. The UpSet plot shows the intersections of metabolites associated with UPF intake (% energy) after multiple testing was corrected for using the Benjamini-Hochberg method; statistical significance was defined as a corrected *p*-value of <0.01. Vertical bars represent the count of shared metabolites across these biospecimen types, with horizontal bars reflecting the total count of metabolites identified in each biospecimen type. The matrix at the bottom indicates which biospecimen types are included in each intersection.

### Pathway analysis

Seven metabolic pathways were associated with UPF intake and common to all three biospecimen types; these include benzoate metabolism; tocopherol metabolism; fatty acid, dicarboxylate metabolism; food component/plant metabolism; leucine, isoleucine and valine metabolism; urea cycle metabolism; arginine and proline metabolism; and tocopherol metabolism (**Supplementary Table 4**).

### Poly-metabolite score generation using IDATA study

For poly-metabolite scores of UPF intake, we selected 28, 33, and 23 metabolites for serum, 24-hour urine, and FMV urine, respectively, using LASSO regression (**Figure 3 & Supplementary Table 5**). RMSE and MAE vales were similar in testing and training datasets, for each biospecimen type, suggesting that the prediction models were not overfit and performing well; additionally, correlations between the scores and UPF intake were high (≥0.47) in testing and training datasets for each biospecimen type (**Supplementary Table 6**). In serum and 24-hour urine analyses, three common metabolites were selected by LASSO regression ≥60 times out of 100 iterations, making them robust predictors (37) of UPF intake. These include two amino acids (i.e., (S)C(S)S-S-methyl cysteine sulfoxide [r_s_= –0.19 to –0.23] and N2-N5-diacetylornithine [r_s_= –0.26 to –0.27]); one partially characterized molecule (i.e., pentoic acid [r_s_= –0.28 to –0.31]), and one carbohydrate (i.e., N6-carboxymethylysine [r_s_= 0.15 to 0.21]; **Figure 3 & Supplementary Figures 3**).

**Figure 3.**
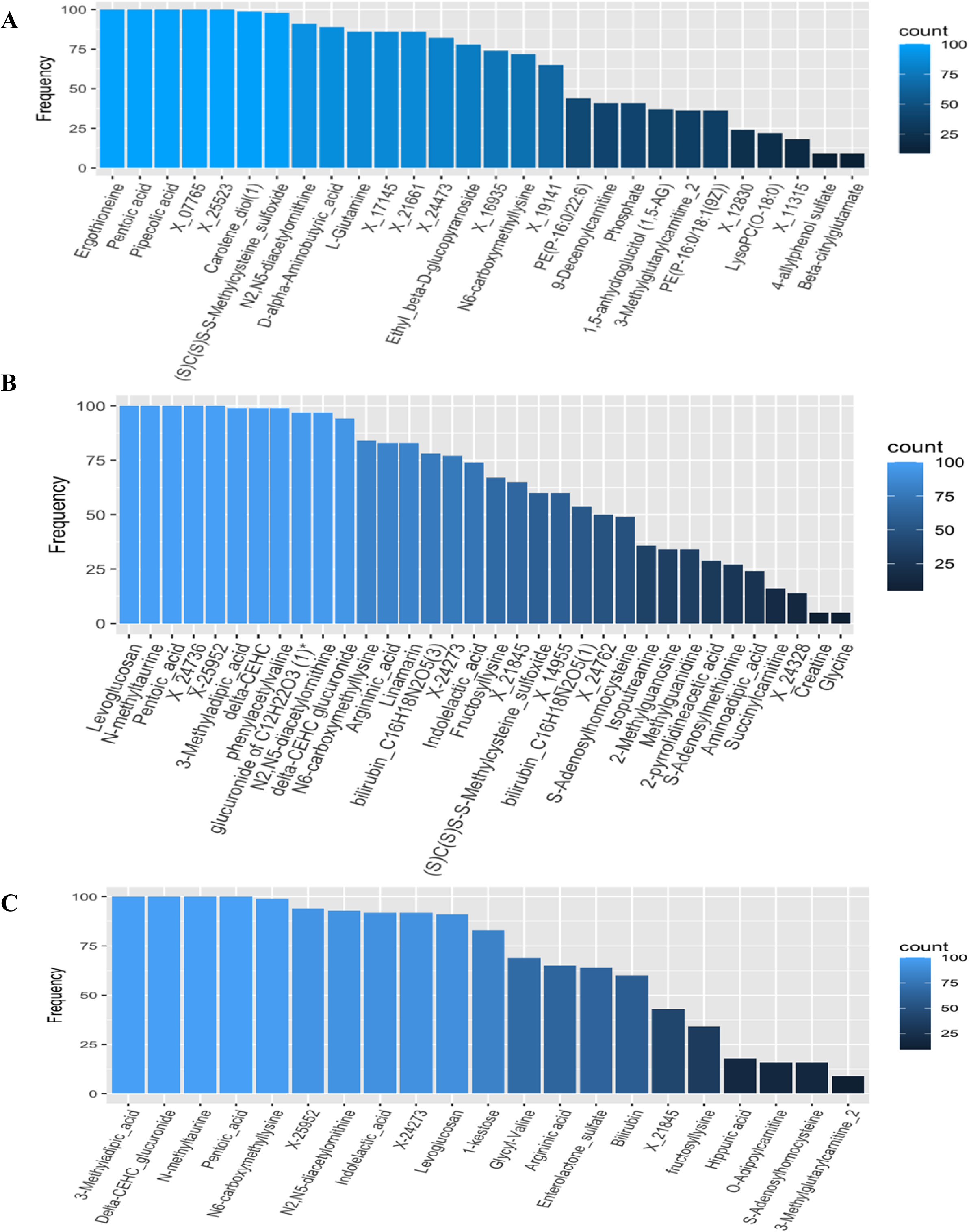
Metabolites selected using LASSO regression as predictive of 12-month average UPF intake (% of energy) in serum (A), 24-hour (B) and FMV (C) urine. This plot shows the number of times a metabolites was selected by the LASSO regression model as predictive of UPF intake (% energy) by biospecimen type. For each biospecimen type, we applied LASSO to randomly selected 80% subsets of the dataset, repeating this procedure 100 times. The y-axis represents the number of times each metabolite was selected out of 100 iterations, and the x-axis displays the names of the LASSO selected metabolites.

### UPF poly-metabolite score applied to a randomized, controlled, crossover-feeding trial

We applied the serum and urine poly-metabolite scores, developed in IDATA, to existing plasma and urine metabolomics data generated in an independent, randomized, controlled, crossover-feeding trial. For the score calculation, 26 out of 28 IDATA-selected serum metabolites were measured in the trial in EDTA plasma as were all selected urine metabolites. We found that the blood– and urine-based poly-metabolite scores differed substantially, within individual, between the 80% and 0% energy from UPF diet phases (all *p*-value for paired t-test<1×10^−5^; **Figures 4**). In contrast, some but not all of the individual metabolites included in the poly-metabolite scores differed, within individual, between diet phases, (**Supplementary Table 7**) suggesting that the score may be better suited as a candidate biomarker of UPF intake than any one individual metabolite. In the subset of four individuals who participated in a second feeding trial of diets with 30% energy from UPF, we found that the poly-metabolite score for 24-hour urine increased stepwise, within individual, with increasing energy from UPF; for EDTA plasma and spot urine ploy-metabolite scores were similar for diets low (i.e., 0 and 30% energy) in UPF and significantly increased for diets high in (80% energy) UPF (**Supplementary Figures 4-6**).

**Figure 4.**
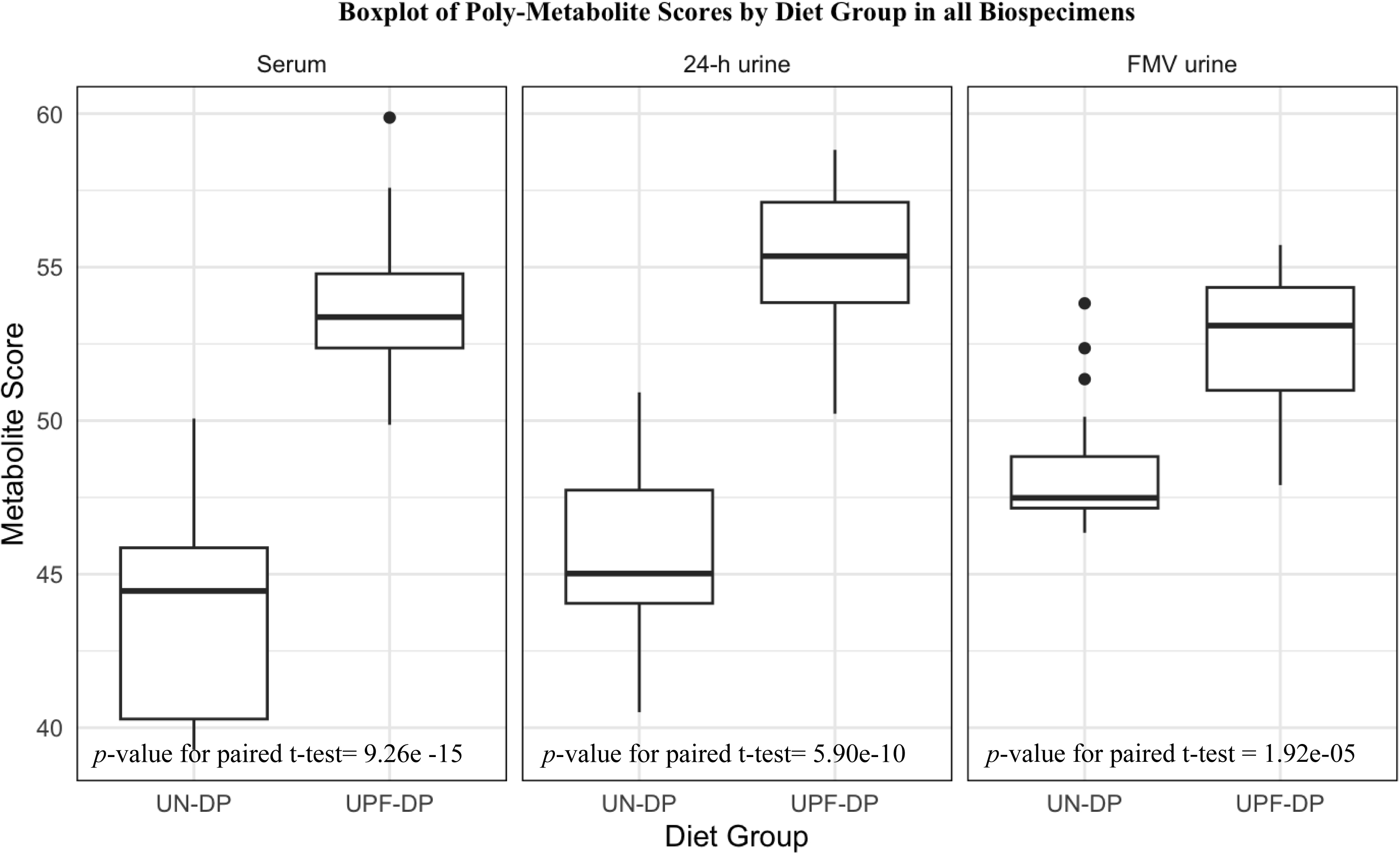
Comparison of poly-metabolite scores for dietary patterns high in ultra-processed food (i.e, 80% energy from UPF) and void of UPF (i.e., 0% energy from UPF) in a randomized, controlled, crossover-feeding trial of domiciled adults (N=20) Box plots show the distribution of poly-metabolite scores, developed in the IDATA study, generated using EDTA plasma and urine metabolomics data from an independent, randomized, controlled, crossover-feeding trial of 20 domiciled healthy participants, who consumed *ad libitum*, for 2 weeks each, an ultra-processed food dietary pattern (i.e., with 80% energy from UPF) and an unprocessed dietary pattern (i.e., 0% energy from UPF). Differences in poly-metabolite scores, within individual, between the two diet phases of the trial were tested using paired t-tests.

## Discussion

Understanding how metabolites are associated with UPF intake and identifying poly-metabolite scores predictive of UPF intake in free-living individuals with varying diets has the potential to complement, extend, and advance research on UPF intake and human health. In the IDATA Study, we identified metabolites significantly correlated with UPF intake using repeated measures of metabolites and dietary intake, assessed via multiple ASA-24s, in 718 generally healthy adults. Moreover, we developed blood and urine poly-metabolite scores for UPF intake, using untargeted serum, 24-hour urine, and FMV urine metabolomics data and a rigorous statistical approach, including training and testing datasets, to ensure the robustness of our results. We then applied these scores to EDTA plasma, 24-hour urine, and spot urine metabolomics data, respectively, that had been previously generated in an independent, randomized, controlled, crossover, feeding trial. We demonstrated the validity of these poly-metabolite scores by showing that they differed, within individual, between consumption of diets high in (80% energy) versus to void of UPF (0% energy). Overall, we identified hundreds of metabolites in serum (n=187) and urine (n=284 in 24-hour urine & n=234 in FMV) that were significantly correlated with UPF intake. These UPF-correlated metabolites are involved in numerous and diverse biological pathways, underscoring the complex impact of diet on the metabolome. For instance, among the metabolites that were positively correlated with UPF intake, there were acylcarnitine derivatives, such as 9-decenoylcarnitine, and sphingolipids, such as the ceramides d18:1/18:0 and d18:1/20:0. Higher serum levels of acylcarnitine have been found in meat eaters compared to vegetarians (22, 38, 39). In contrast, lower levels of the ceramides, d18:1/18:0 and d18:1/20:0, have been associated with better adherence to healthy dietary patterns (40). An even larger number of metabolites were negatively correlated with UPF intake, including beta-cryptoxanthin, which has been identified as a candidate biomarker of fruit and vegetable consumption (22, 41). Thus, the metabolomic profiles that we identified likely reflect a dietary pattern characterized not only by high UPF consumption but also by low whole food, including fresh fruits and vegetables, consumption.

Using LASSO regression models, we identified a set of circulating and urinary metabolites that may serve collectively as predictive biomarkers for UPF intake. In total, we selected 28 serum, 33 24-hour urine, and 23 FMV urine metabolites as predictors of UPF intake. Of these metabolites, three were robust (selected ≥ 60 times out of 100 iterations) biomarkers and were common to both serum and 24-hour urine including two amino acids (i.e., (S)C(S)S-S-Methylcysteine sulfoxide and N2-N5-diacetylornithine) which were both negatively correlated with UPF intake, and a carbohydrate (i.e., N6-carboxymethylysine), which was positively correlated with UPF intake. (S)C(S)S-S-Methylcysteine sulfoxide has been identified as a biomarker of cruciferous vegetable intake (42) indicating that dietary patterns high in UPF intake tend to be low in cruciferous vegetable intake. Serum concentrations of N6-carboxymethylysine, an advanced glycation end-product, have been associated with diabetes incidence and severity as well as with risk of other cardiometabolic diseases (43–45). The positive correlation between N6-carboxymethylysine and UPF intake in our study suggests that a dietary pattern high in UPF may, in part, increase risk of developing type 2 diabetes (46) through this metabolite or related metabolic pathways. This aligns with a systematic umbrella review of existing meta-analyses, which reported that greater exposure to UPF is associated with a higher risk of adverse health outcomes, including type 2 diabetes (12). Moreover, this suggests the utility in using nutritional metabolomics not only to improve exposure measures but also to provide novel insight into biological mechanisms underlying potential UPF-disease associations.

In the serum set, three metabolites in the chemical class, cofactors and vitamins, were negatively correlated with UPF intake; β-cryptoxanthin and carotene diol (1) & (2) are provitamin A carotenoids found in citrus fruits and some vegetables, such as red peppers, that have been identified as candidate biomarkers of fruit and vegetable intake (22, 41). β-cryptoxanthin has also been associated with adherence to a healthy dietary pattern (22, 41, 47). In line with these metabolite findings, in IDATA, we observed that average daily dietary intake of key vitamins and minerals was lower among those with higher UPF intake. This observation aligns with prior research indicating an inverse association between UPF intake and diet quality among both children and adults (48).

The urine metabolites, selected by LASSO regression models, similarly included a diverse array of compounds. Notably, the exogenously derived metabolite, levoglucosan, which was positively correlated with the UPF intake in our study, is a combustion breakdown product of cellulose, which is used as a biopolymer in the food packaging industry (49), and is excreted unmetabolized in urine after exposure (50). This finding supports claims that higher intake of UPF exposes individuals to food contact substances, including some from packaging materials. It is, however, important to note that this dimension of UPF exposure was reflected in urinary and not circulating metabolites. This is an important consideration for future researchers and highlights the need for dietary biomarker discovery not only in blood but also in urine and stool.

The urine metabolites that were selected by LASSO regression were similar to those selected for 24-hour urine but more limited in number. Overall, 203 UPF-metabolite correlations, which were generally similar in both magnitude and direction, were observed in both urine types demonstrating that FMV metabolite-UPF correlations are reflective of those for 24-hour urine. This is important since 24-hour collections are not feasible in most large epidemiological studies. We also show that while there is some overlap between blood and urine for UPF-metabolite correlates and related scores, most are unique to either blood or urine. Distinct metabolites and metabolomic profiles by biospecimen type may also translate to differences in mechanistic insight into potential UPF-health associations in etiologic studies.

Our IDATA findings are consistent with a recent short-term randomized, controlled, crossover, domiciled feeding trial (5), which found that hundreds of circulating (257 out of 993) and urine metabolites (606 out of 1279) differed within individuals between dietary patterns containing 80% UPF and 0% UPF in post-intervention samples. We found that a majority of the metabolites identified as potential candidate biomarker in the feeding trial (i.e., 198 out of 257 circulating metabolites and 341 out of 606 urine metabolites) were also detected in IDATA samples. Of the 59 and 129 serum and urine metabolites that were statistically significant in the IDATA 85% and 81% were associated with UPF intake in the same direction (**Supplementary Tables 8-9**). Three metabolites (i.e., N6-carboxymethylysine, (S)C(S)S-S-Methylcysteine sulfoxide, and pentoic acid) that were selected as robust predictors of UPF intake in both serum and urine were among those reported in the feeding trial. Unsurprisingly, when we tested our IDATA-derived poly-metabolite scores for UPF intake in this same feeding trial, we found that they could discriminate between dietary patterns containing 80% UPF vs 0% UPF, within individual. Thus, our novel poly-metabolite scores could be used as an objective and complementary measure of UPF intake in future epidemiological investigations.

Our study has notable strengths, which contribute to its scientific rigor. Firstly, diet over 12 months was assessed via multiple validated 24-hour dietary recalls, with more than 97% of participants completing four or more. This helped minimize measurement error due to random within-person day-to-day variation in dietary intake, and it also allowed for more accurate and detailed dietary data collection than a food frequency questionnaire. Secondly, we measured more than a thousand metabolites, related to wide range of metabolic pathways, in serial serum and urine samples collected from the same individuals 6-months apart. Serial metabolite measurements again helped to minimize measurement error owing to within-person variability in metabolite measures over time. Thirdly, we not only successfully replicated most of the associations between UPF and metabolites identified in a prior feeding trial, but also demonstrated that our poly-metabolite scores effectively differentiate, within individuals, between diets high in UPF (80%) and diets with 0% UPF in that same trial. While our study population lacked diversity, the study participants, consumed a range of diets, and diets high in UPF were not all the same, suggesting our findings may be generalizable to other populations. Still more research is needed to understand the population specificity of UPF dietary patterns and associated metabolomic profiles.

## Conclusion

In summary, our findings demonstrate that dietary patterns higher in UPFs are associated with distinctive metabolomic profiles in both serum and urine in older, mostly white adults. Further research is needed to understand the full spectrum of metabolites and pathways affected by UPF consumption and to validate our UPF poly-metabolite scores in diverse populations with diverse diets. Finally, research exploring the prospective association between UPF-correlated metabolites and poly-metabolite scores with disease risk is warranted.

## Data Availability

All relevant data are within the manuscript and its Supporting Information files.

## Supplementary Figures

**Figure S1.**
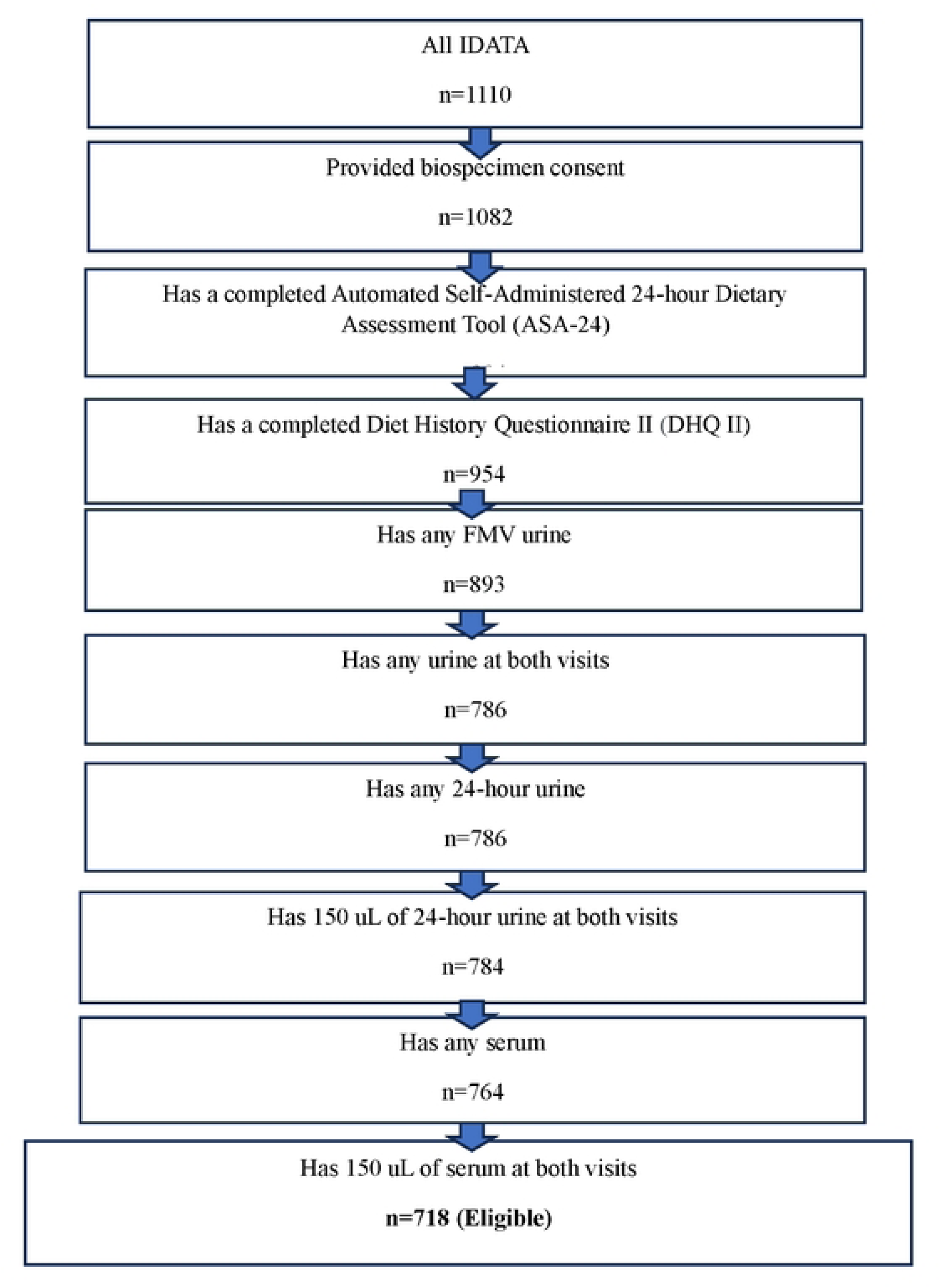
Flowchart for selection of the longitudinal Interactive Diet and Activity Tracking in AARP Study (IDATA) analytic sample for metabolomics analysis.

**Figure S2.**
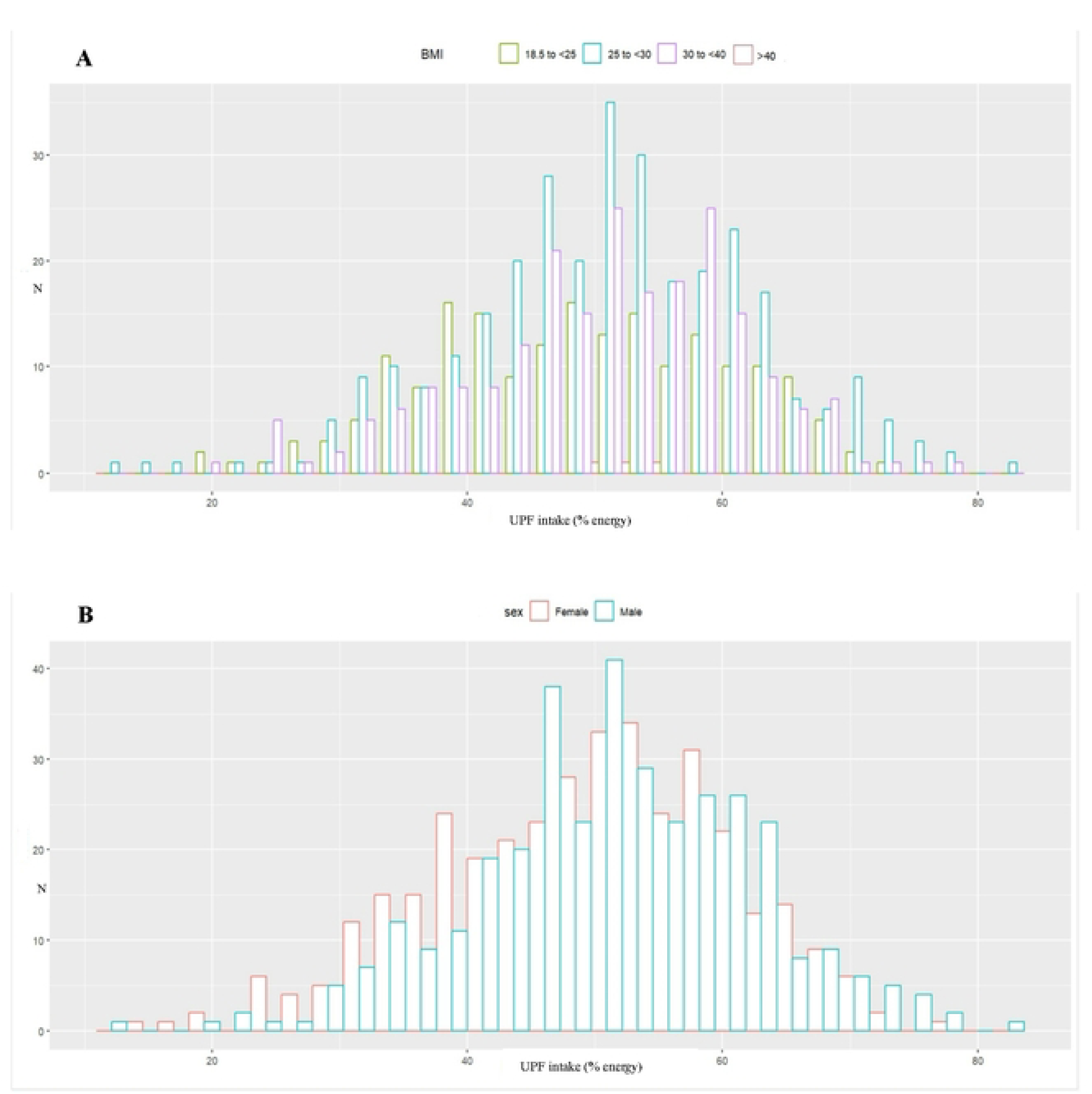
Histogram of ultra-processed food (UPF) intake (% energy) by BMI and sex categories.

**Figure S3.**
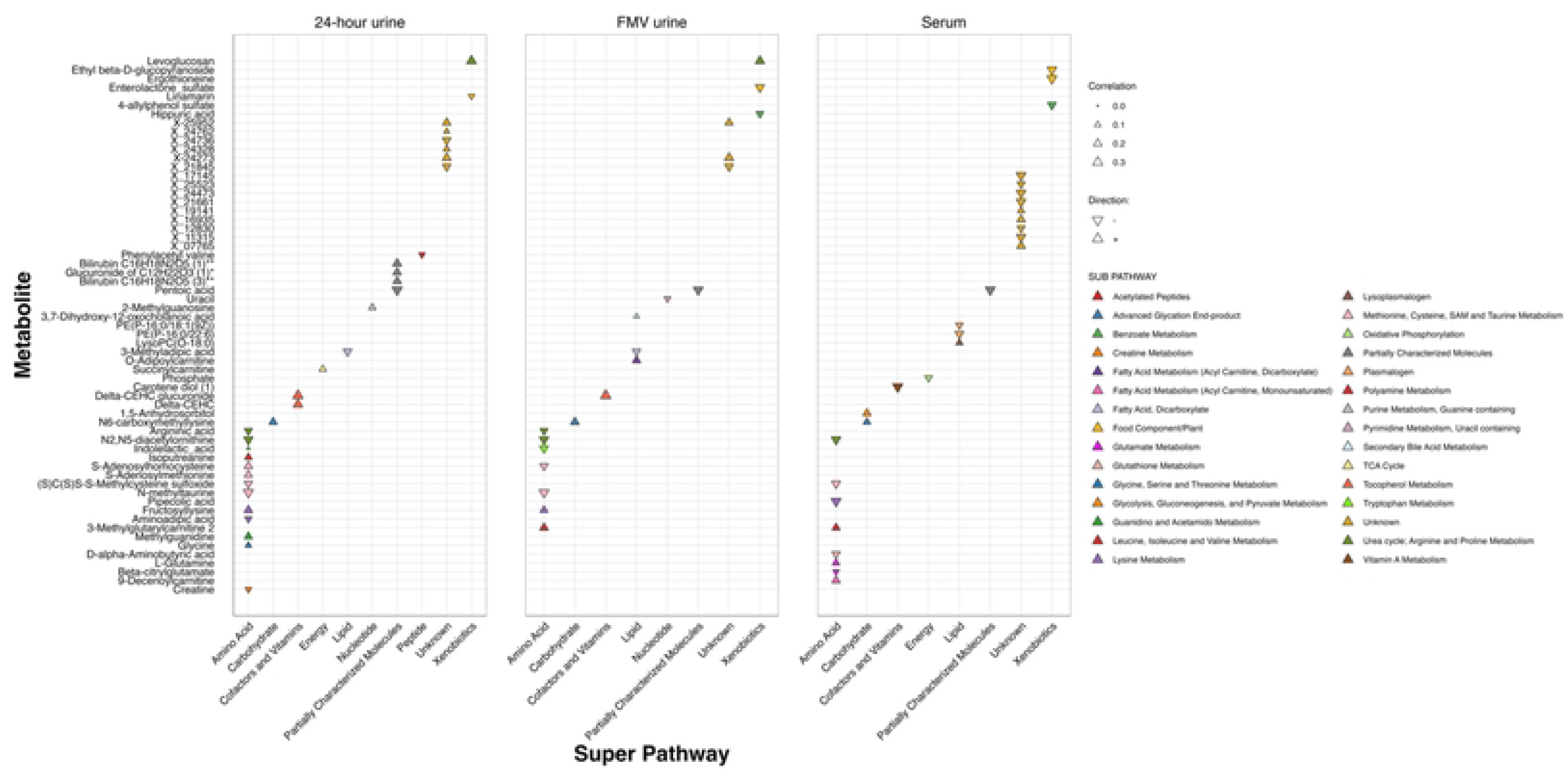
Correlations between ultra-processed food (UPF) intake and metabolites, defined as 12-month mean percentage of total daily energy as measured by multiple 24-hour dietary recalls, for serum, urine (24-hour and FMV) metabolites selected by Least Absolute Shrinkage and Selection Operator (LASSO) regression as predictive of UPF by chemical class.

**Figure S4.**
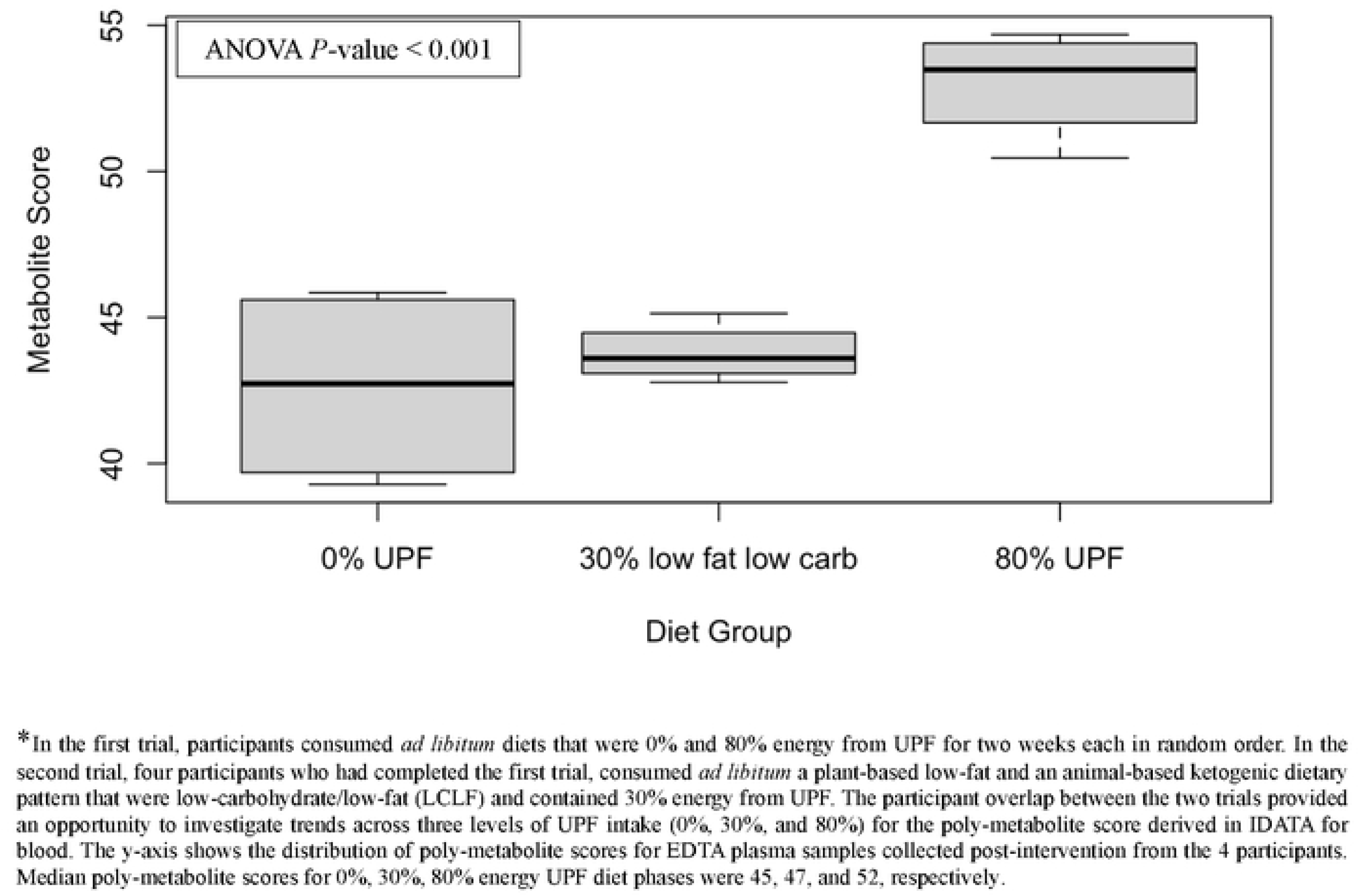
Box plot of poly-metabolite scores, derived in IDATA using serum metabolomics data, and applied to plasma metabolomics data from 4 individuals who pa1ticipated in two distinct randomized, controlled, crossover-feeding trial, of diets that were 0%, 30%, and 80% energy from ultra-processed food (UPF)*

**Figure S5.**
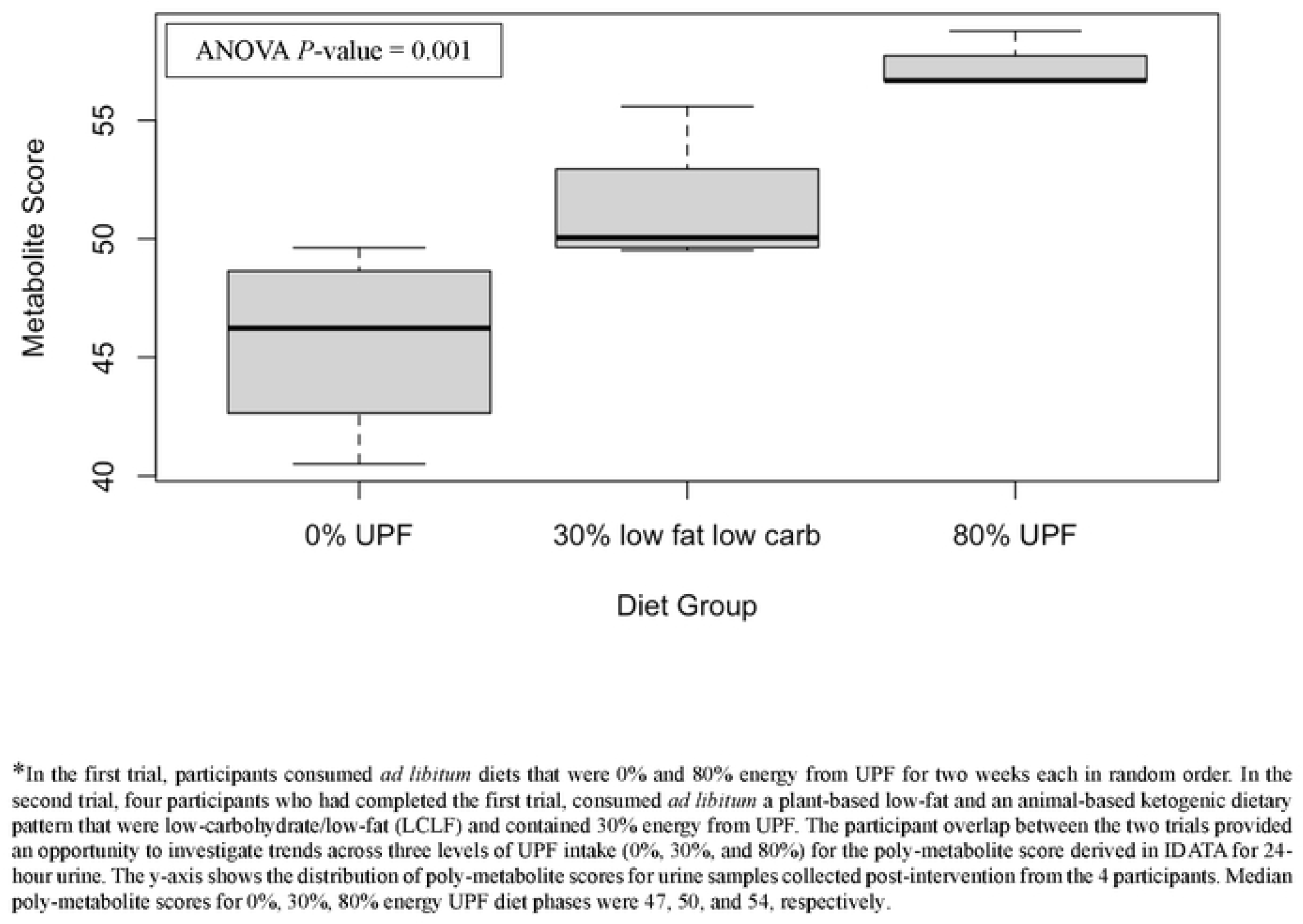
Box plot of poly-metabolite scores, derived in IDATA using 24-hour urine metabolomics data, and applied to urine metabolomics data from 4 individuals who participated in two distinct randomized, controlled, crossover-feeding trial, of diets that were 0%, 30%, and 80% energy from ultra-processed food (UPF)*

**Figure S6.**
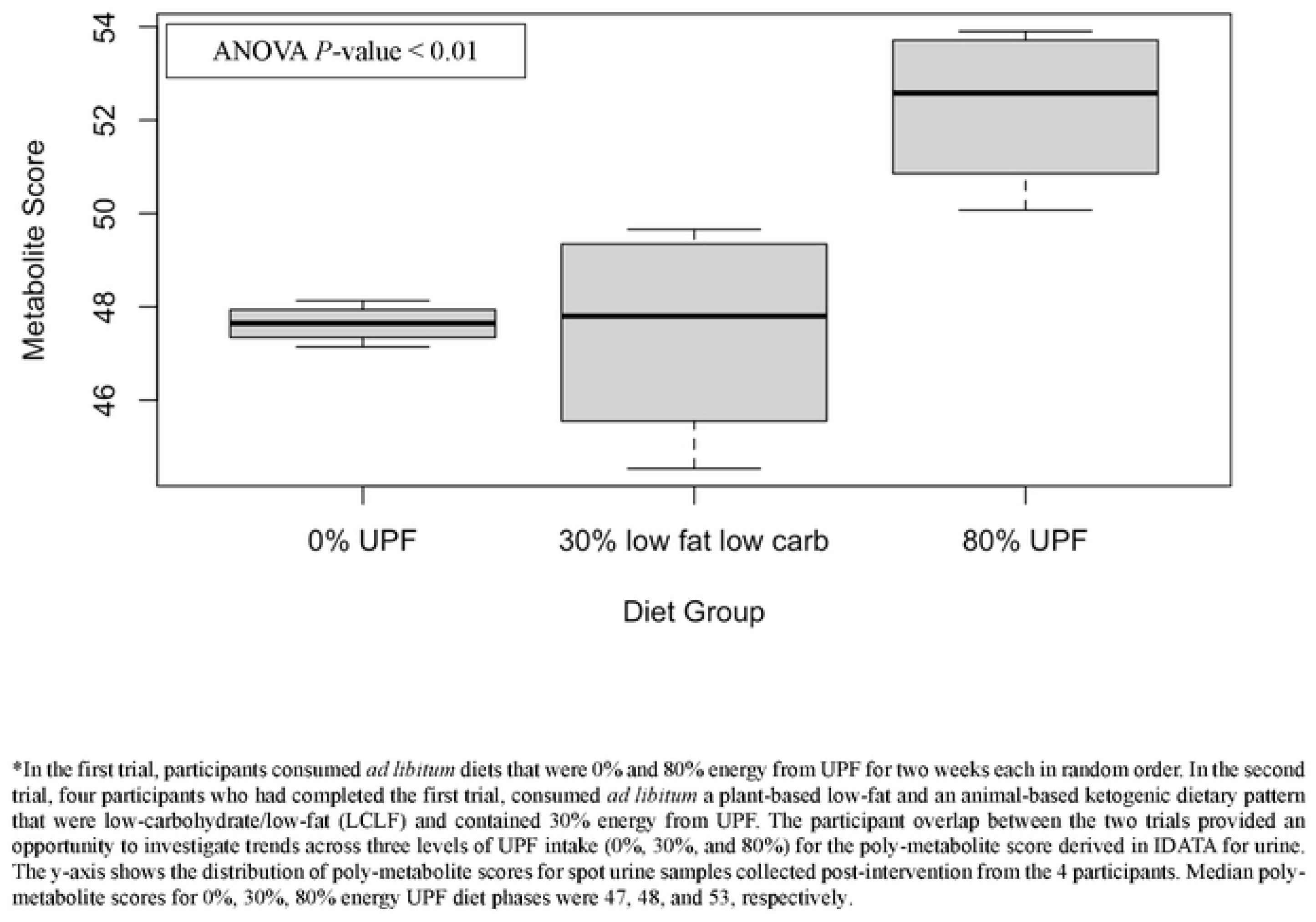
Box plot of poly-metabolite scores, derived in IDATA using FMY urine metabolomics data, and applied to spot urine metabolomics data from 4 individuals who participated in two distinct randomized, controlled, crossover-feeding trial, of diets that were 0%, 30%, and 80% energy from ultra-processed food (UPF)*

## Notes

### Competing Interest Statement

The authors have declared no competing interest.

### Author Declarations

The IDATA study was approved by the National Cancer Institute (NCI) Special Studies Institutional Review Board and is registered on ClinicalTrials.gov (Identifier: NCT03268577); all participants signed informed consent.

## References

1. Mendonça RD, Pimenta AM, Gea A, de la Fuente-Arrillaga C, Martinez-Gonzalez MA, Lopes AC, Bes-Rastrollo M. Ultraprocessed food consumption and risk of overweight and obesity: the University of Navarra Follow-Up (SUN) cohort study. Am J Clin Nutr. 2016;104(5):1433–40.

2. Monteiro CA, Cannon G, Levy RB, Moubarac JC, Louzada ML, Rauber F, et al. Ultra-processed foods: what they are and how to identify them. Public Health Nutr. 2019;22(5):936–41.

3. Baker P, Machado P, Santos T, Sievert K, Backholer K, Hadjikakou M, et al. Ultra-processed foods and the nutrition transition: Global, regional and national trends, food systems transformations and political economy drivers. Obes Rev. 2020;21(12):e13126.

4. Wang L, Martínez Steele E, Du M, Pomeranz JL, O’Connor LE, Herrick KA, et al. Trends in Consumption of Ultraprocessed Foods Among US Youths Aged 2-19 Years, 1999-2018. JAMA. 2021;326(6):519–30.

5. O’Connor LE, Hall KD, Herrick KA, Reedy J, Chung ST, Stagliano M, et al. Metabolomic Profiling of an Ultraprocessed Dietary Pattern in a Domiciled Randomized Controlled Crossover Feeding Trial. J Nutr. 2023;153(8):2181–92.

6. Juul F, Parekh N, Martinez-Steele E, Monteiro CA, Chang VW. Ultra-processed food consumption among US adults from 2001 to 2018. The American Journal of Clinical Nutrition. 2022;115(1):211–21.

7. Beslay M, Srour B, Méjean C, Allès B, Fiolet T, Debras C, et al. Ultra-processed food intake in association with BMI change and risk of overweight and obesity: A prospective analysis of the French NutriNet-Santé cohort. PLoS Med. 2020;17(8):e1003256.

8. Lane MM, Davis JA, Beattie S, Gómez-Donoso C, Loughman A, O’Neil A, et al. Ultraprocessed food and chronic noncommunicable diseases: A systematic review and meta-analysis of 43 observational studies. Obes Rev. 2021;22(3):e13146.

9. Juul F, Martinez-Steele E, Parekh N, Monteiro CA, Chang VW. Ultra-processed food consumption and excess weight among US adults. Br J Nutr. 2018;120(1):90–100.

10. Cordova R, Viallon V, Fontvieille E, Peruchet-Noray L, Jansana A, Wagner KH, et al. Consumption of ultra-processed foods and risk of multimorbidity of cancer and cardiometabolic diseases: a multinational cohort study. Lancet Reg Health Eur. 2023;35:100771.

11. Srour B, Kordahi MC, Bonazzi E, Deschasaux-Tanguy M, Touvier M, Chassaing B. Ultra-processed foods and human health: from epidemiological evidence to mechanistic insights. The Lancet Gastroenterology & Hepatology. 2022;7(12):1128–40.

12. Lane MM, Gamage E, Du S, Ashtree DN, McGuinness AJ, Gauci S, et al. Ultra-processed food exposure and adverse health outcomes: umbrella review of epidemiological meta-analyses. BMJ. 2024;384:e077310.

13. de Araújo TP, de Moraes MM, Afonso C, Santos C, Rodrigues SSP. Food Processing: Comparison of Different Food Classification Systems. Nutrients. 2022;14(4):729.

14. Braesco V, Souchon I, Sauvant P, Haurogné T, Maillot M, Féart C, Darmon N. Ultra-processed foods: how functional is the NOVA system? European Journal of Clinical Nutrition. 2022;76(9):1245–53.

15. Drewnowski A, Detzel P, Klassen-Wigger P. Perspective: Achieving Sustainable Healthy Diets Through Formulation and Processing of Foods. Current Developments in Nutrition. 2022;6(6):nzac089.

16. Monteiro CA, Cannon G, Moubarac JC, Levy RB, Louzada MLC, Jaime PC. The UN Decade of Nutrition, the NOVA food classification and the trouble with ultra-processing. Public Health Nutr. 2018;21(1):5–17.

17. Freedman LS, Midthune D, Carroll RJ, Tasevska N, Schatzkin A, Mares J, et al. Using regression calibration equations that combine self-reported intake and biomarker measures to obtain unbiased estimates and more powerful tests of dietary associations. Am J Epidemiol. 2011;174(11):1238–45.

18. Brouwer-Brolsma EM, Brennan L, Drevon CA, van Kranen H, Manach C, Dragsted LO, et al. Combining traditional dietary assessment methods with novel metabolomics techniques: present efforts by the Food Biomarker Alliance. Proc Nutr Soc. 2017;76(4):619–27.

19. Landberg R, Karra P, Hoobler R, Loftfield E, Huybrechts I, Rattner JI, et al. Dietary biomarkers-an update on their validity and applicability in epidemiological studies. Nutr Rev. 2023.

20. Playdon MC, Tinker LF, Prentice RL, Loftfield E, Hayden KM, Van Horn L, et al. Measuring diet by metabolomics: a 14-d controlled feeding study of weighed food intake. Am J Clin Nutr. 2023.

21. Brennan L, Hu FB, Sun Q. Metabolomics Meets Nutritional Epidemiology: Harnessing the Potential in Metabolomics Data. Metabolites. 2021;11(10):709.

22. Kim H, Rebholz CM. Metabolomic Biomarkers of Healthy Dietary Patterns and Cardiovascular Outcomes. Current Atherosclerosis Reports. 2021;23(6):26.

23. Mazzilli KM, McClain KM, Lipworth L, Playdon MC, Sampson JN, Clish CB, et al. Identification of 102 Correlations between Serum Metabolites and Habitual Diet in a Metabolomics Study of the Prostate, Lung, Colorectal, and Ovarian Cancer Trial. The Journal of Nutrition. 2020;150(4):694–703.

24. Su D, Chen J, Du S, Kim H, Yu B, Wong KE, et al. Metabolomic Markers of Ultra-Processed Food and Incident CKD. Clinical Journal of the American Society of Nephrology. 2023;18(3):327–36.

25. Handakas E, Chang K, Khandpur N, Vamos EP, Millett C, Sassi F, et al. Metabolic profiles of ultra-processed food consumption and their role in obesity risk in British children. Clinical Nutrition. 2022;41(11):2537–48.

26. Subar AF, Potischman N, Dodd KW, Thompson FE, Baer DJ, Schoeller DA, et al. Performance and Feasibility of Recalls Completed Using the Automated Self-Administered 24-Hour Dietary Assessment Tool in Relation to Other Self-Report Tools and Biomarkers in the Interactive Diet and Activity Tracking in AARP (IDATA) Study. Journal of the Academy of Nutrition and Dietetics. 2020;120(11):1805–20.

27. National Cancer Institute. Automated Self-Administered 24-Hour (ASA24) Dietary Assessment Tool. Available from: https://epi.grants.cancer.gov/asa24/ [Accessed March15, 2024].

28. Park Y, Dodd KW, Kipnis V, Thompson FE, Potischman N, Schoeller DA, et al. Comparison of self-reported dietary intakes from the Automated Self-Administered 24-h recall, 4-d food records, and food-frequency questionnaires against recovery biomarkers. Am J Clin Nutr. 2018;107(1):80–93.

29. Steele EM, O’Connor LE, Juul F, Khandpur N, Galastri Baraldi L, Monteiro CA, et al. Identifying and Estimating Ultraprocessed Food Intake in the US NHANES According to the Nova Classification System of Food Processing. J Nutr. 2023;153(1):225–41.

30. Agriculture ARSUSDo. Food Surveys Research Group: Bestville, MD [Available from: https://www.ars.usda.gov/northeast-area/beltsville-md-bhnrc/beltsville-human-nutrition-research-center/food-surveys-research-group/docs/fndds/

31. Fukagawa NK, McKillop K, Pehrsson PR, Moshfegh A, Harnly J, Finley J. USDA’s FoodData Central: what is it and why is it needed today? Am J Clin Nutr. 2022;115(3):619–24.

32. Martínez Steele E, Juul F, Neri D, Rauber F, Monteiro CA. Dietary share of ultra-processed foods and metabolic syndrome in the US adult population. Prev Med. 2019;125:40–8.

33. Ford L, Mitchell M, Wulff J, Evans A, Kennedy A, Elsea S, et al. Clinical metabolomics for inborn errors of metabolism. Adv Clin Chem. 2022;107:79–138.

34. Sampson JN, Boca SM, Shu XO, Stolzenberg-Solomon RZ, Matthews CE, Hsing AW, et al. Metabolomics in epidemiology: sources of variability in metabolite measurements and implications. Cancer Epidemiol Biomarkers Prev. 2013;22(4):631–40.

35. Fisher RA. Statistical Methods for Research Workers. In: Kotz S, Johnson NL, editors. Breakthroughs in Statistics: Methodology and Distribution. New York, NY: Springer New York; 1992. p. 66-70.

36. Hall KD, Guo J, Courville AB, Boring J, Brychta R, Chen KY, et al. Effect of a plant-based, low-fat diet versus an animal-based, ketogenic diet on ad libitum energy intake. Nat Med. 2021;27(2):344–53.

37. Meinshausen N, Bühlmann P. Stability Selection. Journal of the Royal Statistical Society Series B: Statistical Methodology. 2010;72(4):417–73.

38. Schmidt JA, Rinaldi S, Ferrari P, Carayol M, Achaintre D, Scalbert A, et al. Metabolic profiles of male meat eaters, fish eaters, vegetarians, and vegans from the EPIC-Oxford cohort. Am J Clin Nutr. 2015;102(6):1518–26.

39. Tong TYN, Koulman A, Griffin JL, Wareham NJ, Forouhi NG, Imamura F. A Combination of Metabolites Predicts Adherence to the Mediterranean Diet Pattern and Its Associations with Insulin Sensitivity and Lipid Homeostasis in the General Population: The Fenland Study, United Kingdom. J Nutr. 2020;150(3):568–78.

40. Tanaka T, Talegawkar SA, Jin Y, Candia J, Tian Q, Moaddel R, et al. Metabolomic Profile of Different Dietary Patterns and Their Association with Frailty Index in Community-Dwelling Older Men and Women. Nutrients. 2022;14(11).

41. Rebholz CM, Lichtenstein AH, Zheng Z, Appel LJ, Coresh J. Serum untargeted metabolomic profile of the Dietary Approaches to Stop Hypertension (DASH) dietary pattern. Am J Clin Nutr. 2018;108(2):243–55.

42. Edmands WM, Beckonert OP, Stella C, Campbell A, Lake BG, Lindon JC, et al. Identification of human urinary biomarkers of cruciferous vegetable consumption by metabonomic profiling. J Proteome Res. 2011;10(10):4513–21.

43. Luft VC, Duncan BB, Schmidt MI, Chambless LE, Pankow JS, Hoogeveen RC, et al. Carboxymethyl lysine, an advanced glycation end product, and incident diabetes: a case-cohort analysis of the ARIC Study. Diabet Med. 2016;33(10):1392–8.

44. Singh R, Barden A, Mori T, Beilin L. Advanced glycation end-products: a review. Diabetologia. 2001;44(2):129–46.

45. Vlassara H, Striker GE. Advanced glycation endproducts in diabetes and diabetic complications. Endocrinol Metab Clin North Am. 2013;42(4):697–719.

46. Duan M, Vinke PC, Navis GJ, Corpeleijn E, Dekker LH. Associations of ultra-processed food and its consumption patterns with incident type 2 diabetes. European Journal of Public Health. 2020;30(Supplement_5).

47. McCullough ML, Maliniak ML, Stevens VL, Carter BD, Hodge RA, Wang Y. Metabolomic markers of healthy dietary patterns in US postmenopausal women. Am J Clin Nutr. 2019;109(5):1439–51.

48. Liu J, Steele EM, Li Y, Karageorgou D, Micha R, Monteiro CA, Mozaffarian D. Consumption of Ultraprocessed Foods and Diet Quality Among U.S. Children and Adults. Am J Prev Med. 2022;62(2):252–64.

49. Perera KY, Jaiswal AK, Jaiswal S. Biopolymer-Based Sustainable Food Packaging Materials: Challenges, Solutions, and Applications. Foods. 2023;12(12).

50. Naeher LP, Barr DB, Adetona O, Simpson CD. Urinary levoglucosan as a biomarker for woodsmoke exposure in wildland firefighters. Int J Occup Environ Health. 2013;19(4):304–10.

